# SARS-CoV-2 Testing Disparities in Massachusetts

**DOI:** 10.1101/2020.11.02.20224469

**Authors:** Scott Dryden-Peterson, Gustavo E. Velásquez, Thomas J. Stopka, Sonya Davey, Shahin Lockman, Bisola Ojikutu

## Abstract

**Objective:** Early deficiencies in testing capacity contributed to poor control of transmission of severe acute respiratory syndrome coronavirus 2 (SARS-CoV-2). In the context of marked improvement in SARS-CoV-2 testing infrastructure, we sought to examine the alignment of testing with epidemic intensity to mitigate subsequent waves of COVID-19 in Massachusetts.

**Methods:** We compiled publicly available weekly SARS-CoV-2 molecular testing data for period (May 27 to October 14, 2020) following the initial COVID-19 wave. We defined testing intensity as weekly SARS-CoV-2 tests performed per 100,000 population and used weekly test positivity (percent of tests positive) as a measure of epidemic intensity. We considered optimal alignment of testing resources to be matching community ranks of testing and positivity. In communities with a lower rank of testing than positivity in a given week, the testing gap was calculated as the additional tests required to achieve matching ranks. Multivariable Poisson modeling was utilized to assess for trends and association with community characteristics.

**Results:** During the observation period, 4,262,000 tests were reported in Massachusetts and the misalignment of testing with epidemic intensity increased. The weekly testing gap increased 9.0% per week (adjusted rate ratio [aRR]: 1.090, 95% confidence interval [CI]: 1.08-1.10). Increasing levels of community socioeconomic vulnerability (aRR: 1.35 per quartile increase, 95% CI: 1.23-1.50) and the highest quartile of minority and language vulnerability (aRR: 1.46, 95% CI 0.96-1.49) were associated with increased testing gaps, but the latter association was not statistically significant. Presence of large university student population (>10% of population) was associated with a marked decrease in testing gap (aRR 0.21, 95% CI: 0.12-0.38).

**Conclusion:** These analyses indicate that despite objectives to promote equity and enhance epidemic control in vulnerable communities, testing resources across Massachusetts have been disproportionally allocated to more affluent communities. Worsening structural inequities in access to SARS-CoV-2 testing increase the risk for another intense wave of COVID-19 in Massachusetts, particularly among vulnerable communities.

## SARS-CoV-2 Testing Disparities in Massachusetts

Early deficiencies in testing capacity contributed to poor control of transmission of severe acute respiratory syndrome coronavirus 2 (SARS-CoV-2),^1^ particularly among minority and socioeconomically vulnerable communities.^2,3^ Allocating testing resources to locations of greatest need are important to mitigate subsequent waves of COVID-19.^4^ In the context of marked improvement in SARS-CoV-2 testing infrastructure, we sought to examine the alignment of testing with epidemic intensity in Massachusetts.

## Methods

We compiled publicly available weekly SARS-CoV-2 molecular testing data from the Massachusetts Department of Public Health (MDPH) and Boston Public Health Commission (BPHC) for period (May 27 to October 14, 2020) following the initial COVID-19 wave. The BPHC reported tests of unique Boston residents whereas MDPH reported total tests of residents across the state, including repeat testing of individuals. Consequently, we performed separate analyses for all residents of Massachusetts (351 cities and towns) and Boston residents alone (15 neighborhoods).

We defined testing intensity as weekly SARS-CoV-2 tests performed per 100,000 population and used weekly test positivity (percent of tests positive) as a measure of epidemic intensity. We considered optimal alignment of testing resources to be matching community ranks of testing and positivity. In communities with a lower rank of testing than positivity in a given week, the testing gap was calculated as the additional tests required to achieve matching ranks. Communities with matching or higher ranks of testing compared with positivity were considered to have no testing gap.

Data from the American Community Survey (2014-2018) were used to characterize communities. Negative binomial Poisson models with robust sandwich estimators were fit to assess trends in the magnitude of the testing gap over time and associations with selected Social Vulnerability Index domains (Socioeconomic Status, and Minority Status and Language), and large university student population (>10% of residents). Due to collinearity, the model of Boston neighborhoods only assessed associations with time and socioeconomic vulnerability.

## Results

During the observation period, 4,262,000 tests were reported in Massachusetts and 44,180 detected SARS-CoV-2 (1.0% positivity). Median cumulative COVID-19 incidence among communities was 339 (range 0-6670) per 100,000. Median testing intensity across communities was 41,000 (range 5350-274,000) per 100,000 with observed increased testing in the Boston metropolitan area, vacation communities, and university towns (**Figure 1**).

**Figure 1.**
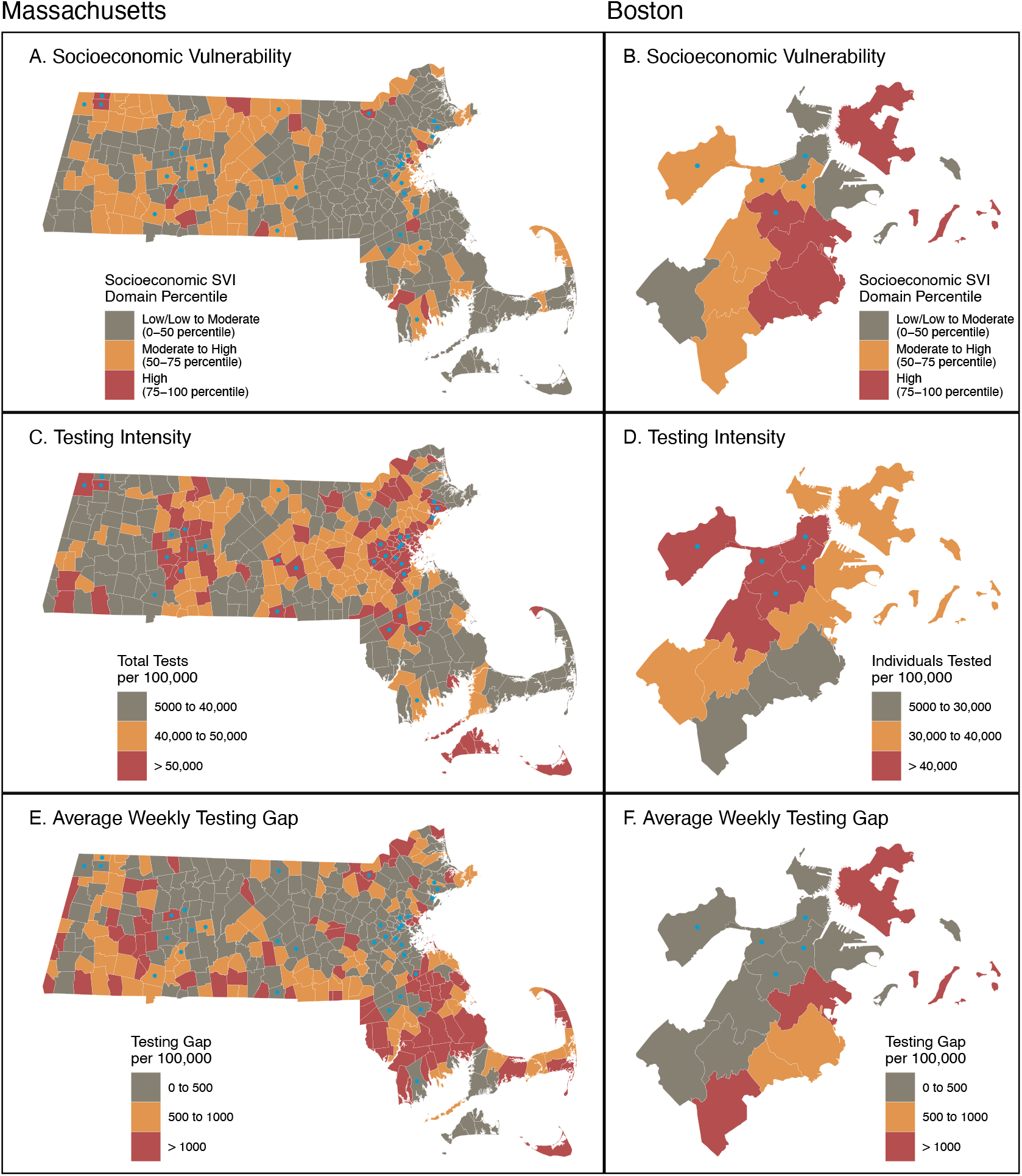
Socioeconomic vulnerability, SARS-CoV-2 testing intensity, and mean SARS-CoV-2 testing gap among Massachusetts cities and towns, and Boston neighborhoods, May 27-October 14, 2020. Community socioeconomic vulnerability (A, B) estimated by the percentile from the Socioeconomic Status domain of the Social Vulnerability Index (SVI, Centers for Disease Control and Prevention). Testing intensity (C, D) includes total tests (including repeat tests in same individual) for Massachusetts but tested individuals (not including repeat testing) for Boston neighborhoods. Average weekly testing gap (E, F) calculated as mean gap (number of additional tests needed so that community rank of testing would match rank of positivity) during the observation period. Blue colored circles indicate communities with large university student populations (>10% of residents). Data broken into three categories for illustrative purposes, but statistical models considered gap as continuous and socioeconomic vulnerability as quartiles of the United States population.

In multivariable model of statewide testing, the misalignment of testing with epidemic intensity grew, with the testing gap increasing 9.0% per week (adjusted rate ratio [aRR]: 1.090, 95% confidence interval [CI]: 1.08-1.10, p<0.001) (**Figure 2**). Increasing levels of community socioeconomic vulnerability (aRR: 1.35 per quartile increase, 95% CI: 1.23-1.50, p<0.001) and the highest quartile of minority and language vulnerability (aRR: 1.46, 95% CI 0.96-1.49, p=0.076) were associated with increased testing gaps, but the latter association was not statistically significant. Presence of large university student population was associated with a marked decrease in testing gap (aRR 0.21, 95% CI: 0.12-0.38, p<0.001).

**Figure 2.**
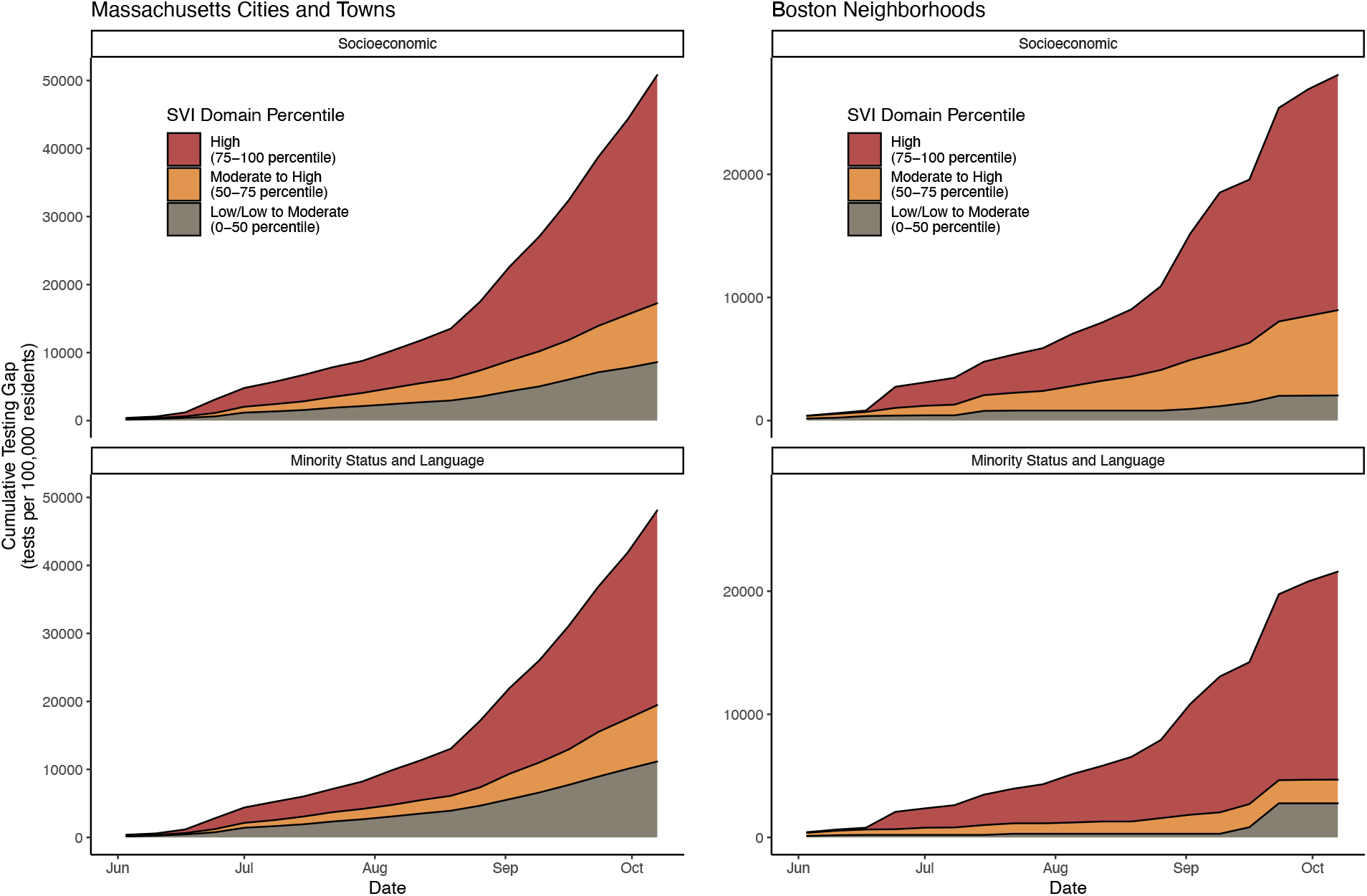
Social vulnerability and relative SARS-CoV-2 testing gap among Massachusetts cities and towns and Boston neighborhoods, May 27 to October 14, 2020. Community social vulnerability was estimated using the Socioeconomic Status and Minority Status and Language domains of the Social Vulnerability Index (SVI, Centers for Disease Control and Prevention) aggregated to the community units used by the Massachusetts Department of Public Health (Massachusetts cities and towns) and the Boston Public Health Commission (Boston neighborhoods). Data broken into three categories of percentiles of the United States population.

Similar findings were observed within Boston, with testing gaps increasing (aRR: 1.08 per week, 95% CI: 1.03-1.13, p=0.003) and larger testing gaps in more socioeconomically vulnerable neighborhoods (aRR 2.51 per quartile increase, 95% CI: 1.56-4.03, p< 0.001).

## Discussion

These analyses indicate that despite objectives to promote equity and enhance epidemic control in vulnerable communities,^5,6^ testing resources across Massachusetts have been disproportionally allocated to more affluent communities. Expanded testing on university campuses contributed to inequity, but disparity persisted following adjustment for these additional tests.

This study has limitations, including use of test positivity to estimate epidemic intensity despite varying rates of asymptomatic testing and aggregation at the community level that could underestimate disparities.

Worsening structural inequities in access to SARS-CoV-2 testing increase the risk for another intense wave of COVID-19 in Massachusetts, particularly among vulnerable communities.

## Data Availability

Data and analysis code shared.

https://github.com/sldrydenpeterson/MA-SARSCoV2-Testing-Alignment.git

